# Derivation and validation of clinical subphenotypes of adult sepsis in Africa: A latent class analysis of infection site and organ dysfunction

**DOI:** 10.64898/2026.09.04.26362265

**Authors:** Phoebe Gruccio, Louisa Edwards, Reagan Kakande, Muhammad Zulfiqar, Michael Ndyomugabe, Victor Adejayan, Philemon Ojuman, William S. Girard, Eva Otoupalova, Paulin R. Banguti, Dennis Hopkinson, Aamer Syed, Jean Paul Mvukiyehe, Venance P. Maro, Matthew P. Rubach, John A. Crump, Cozie Gwaikolo, Richard Ssekitoleko, Elisabeth D. Riviello, Edwin Nuwagira, Christopher C. Moore

## Abstract

**Objective:** Sepsis is a heterogeneous syndrome characterized by substantial variation in infection source, organ dysfunction, and clinical outcomes, and a disproportionate burden in Africa. We aimed to derive and externally validate clinical subphenotypes among adults hospitalized with sepsis in Africa.

**Design:** We used latent class analysis with infection source and organ dysfunction as model inputs. We labeled classes post hoc according to their predominant clinical features. We evaluated associations between subphenotypes and 30-day mortality using multivariable logistic regression.

**Setting:** Hospitals in Uganda, Rwanda, Liberia, Tanzania, and Malawi.

**Patients:** We pooled data from three prospective cohorts of adults hospitalized with sepsis in Uganda, Rwanda, and Liberia for model derivation and independent cohorts from Tanzania and Malawi for external validation.

**Interventions:** None.

**Measurements and Main Results:** The derivation dataset included 467 adults with sepsis, of whom 128 (27.4%) died within 30 days of admission. Latent class analysis identified four subphenotypes: Class 1 (heterogeneous; n=227; 30.8% mortality), Class 2 (abdominal; n=141; 31.9%), Class 3 (pulmonary; n=68; 16.2%), and Class 4 (soft tissue; n=31; 6.5%). Mortality differed significantly across the four classes (p=0.003). Compared with Class 1, Classes 3 (aOR 0.22, 95% CI 0.10–0.47) and 4 (aOR 0.14, 95% CI 0.02–0.52) had lower odds of 30-day mortality. External validation in 316 adults demonstrated high classification certainty (entropy=0.78; mean maximum posterior probabilities, 0.89-0.92), with similar mortality patterns across corresponding subphenotypes.

**Conclusions:** Latent class analysis identified reproducible clinical sepsis subphenotypes defined by infection source and organ dysfunction with distinct mortality risks. These externally validated subphenotypes may improve risk stratification and inform subphenotype-based clinical trials and treatment strategies in Africa.

**KEY POINTS:** *Question:* Can routinely available infection-source and organ-dysfunction data identify reproducible clinical sepsis subphenotypes with distinct mortality risks among adults hospitalized in Africa?

*Findings:* In a pooled cohort study using latent class analysis, we identified four clinical sepsis subphenotypes among 467 adults and externally validated them in 316 adults from independent African cohorts. The subphenotypes had significantly different 30-day mortality, ranging from 6.5% to 31.9%, with similar mortality patterns in the validation cohorts.

*Meaning:* Clinically identifiable sepsis subphenotypes based on routinely available data may provide a pragmatic framework for risk stratification and patient stratification in future sepsis trials in Africa.

## INTRODUCTION

Sepsis occurs due to life-threatening organ dysfunction caused by a dysregulated host response to infection (1). It is a leading cause of global morbidity and mortality, with an estimated 48.9 million cases and 11 million associated deaths annually (2). The burden of sepsis is disproportionately high in Africa, where hospital-based mortality is 30% to 50% (3). Sepsis encompasses a broad spectrum of infections and patterns of organ dysfunction that differ in pathophysiology, clinical presentation, and prognosis (4–6). This heterogeneity complicates risk stratification and clinical management, and may contribute to the limited success of therapeutic strategies that treat sepsis as a single disease entity.

Differences in host responses across infection sites contribute to variation in clinical presentation and prognosis (6). Large multicenter studies have shown that mortality differs by infection source, with higher mortality among patients with intra-abdominal and central nervous system infections than among those with urinary tract infections (4). These observations have prompted efforts to identify distinct sepsis subphenotypes defined by shared clinical, physiologic, biochemical, or biomarker characteristics. In high-income settings, latent class analysis has identified reproducible clinical subphenotypes characterized by distinct patterns of inflammation, organ dysfunction, and outcomes (7). Despite the disproportionate burden of sepsis in Africa, studies characterizing clinical sepsis subphenotypes in this population remain limited (8). Identifying clinically defined subphenotypes in African cohorts may improve risk stratification and guide treatment strategies.

Accordingly, we examined the relationship between infection source, patterns of organ dysfunction, and mortality by 30 days from admission among hospitalized adults with sepsis in Africa. We used latent class analysis to derive and externally validate clinically meaningful sepsis subphenotypes based on infection source and organ dysfunction across multiple African cohorts.

## METHODS

For the derivation dataset, we pooled data from three cohorts of adults with sepsis admitted to the University Teaching Hospitals of Kigali (CHUK) and Butare (CHUB) in Rwanda (n=183; 2017) (9) John F. Kennedy Hospital in Liberia (n=202; 2018; unpublished data), and Mbarara Regional Referral Hospital in Uganda (n=300; 2019-2023) (10). We used the pooled derivation dataset to develop the latent class model. The validation dataset comprised three additional cohorts: St. Joseph’s and Songea Regional Referral Hospitals in Tanzania (n=709; 2006), (11) Kilimanjaro Christian Medical Centre and Mawenzi Regional Referral Hospital in Tanzania (n=403; 2007-2008), (12) and Queen Elizabeth Central Hospital in Malawi (n=361; 2012) (13). We pooled data from these cohorts to form the validation dataset, to which we subsequently applied the latent class model developed in the derivation dataset. All original research involving human participants was approved by the appropriate ethics committees and conducted in accordance with the principles of the Declaration of Helsinki.

We applied the same eligibility criteria to participants in the pooled derivation and validation datasets to define the final analytic cohorts. Inclusion criteria were age ≥18 years, admission to an accident and emergency or medical ward, and sepsis defined as a clinical diagnosis of infection and ≥2 quick Sequential Organ Failure Assessment (qSOFA) criteria: respiratory rate ≥22 breaths per minute, systolic blood pressure ≤100 mmHg, or altered mental status, defined as a Glasgow Coma Scale (GCS) score <15 or other documentation of altered mental status (1).

### Clinical definitions

Based on documented admission and discharge diagnoses, we categorized infections to the following nine infection sites: central nervous system (CNS), pulmonary, abdominal, genitourinary, soft tissue, bacteremia, malaria, tuberculosis, or undifferentiated **(Supplementary Table 1)**. We categorized organ dysfunction into three types: cardiovascular (heart rate ≥120 beats per minute, systolic blood pressure <90 mmHg, mean arterial pressure <65 mmHg, or receipt of a vasopressor), respiratory (respiratory rate ≥30 breaths per minute or oxygen saturation <90%), or neurological (GCS score <15 or other documentation of altered mental status) **(Supplementary table 2)** (14).

### Statistical analysis

#### Clinical variables

We imputed missing data points of vital signs needed to calculate organ dysfunction using k-nearest neighbors (14). We summarized categorical variables as numbers and percentages and compared them using the chi-squared test. We summarized continuous variables as medians with interquartile ranges (IQRs) and compared their distributions between groups with the Mann-Whitney U test. We assessed associations between categorical variables using Cramér’s V, which range from 0 (no association) to 1 (perfect association). We interpreted values of approximately 0.10, 0.30, and 0.50 as weak, moderate, and strong associations, respectively (15, 16). We used logistic regression to examine associations between clinical variables, including the Universal Vital Assessment (UVA) mortality risk score, (14) and mortality by 30 days from admission to evaluate both independent effects and pairwise interactions between infection site and organ dysfunction. We adjusted all models for age and sex, as well as enrollment site (Uganda) in the derivation dataset. We performed all analyses using R (R Foundation for Statistical Computing, Vienna, Austria) and RStudio (version 2025.09.1+401; Posit Software, PBC, Boston, MA, USA).

#### Latent class analysis

To reduce dimensionality and derive clinically meaningful subphenotypes, we performed latent class analysis using the nine infection sites and the three organ dysfunction types as input variables. The analysis was data-driven and did not incorporate prior information about class structure or previously described sepsis subphenotypes. We selected optimal models using Akaike Information Criterion (AIC), Bayesian Information Criterion (BIC), entropy, and clinical interpretability; and prioritized models that balanced goodness of fit, parsimony, clear class separation, and clinical relevance (17). After class assignment, we described the demographic and clinical characteristics, infection sites, organ dysfunctions, and mortality by 30 days from admission of participants within each class. We assigned descriptive labels to the classes post hoc based on their predominant distributions of infection sites and organ dysfunctions.

To validate our findings, we separately assigned participants in the validation dataset to the latent classes identified in the derivation cohort using mean posterior probabilities (17). We assessed the fit of the validation dataset participants in the derived latent classes using mean maximum posterior probabilities. We used multivariable logistic regression to assess the association between latent class membership and mortality by 30 days from admission separately in the derivation and validation datasets. We assessed model discrimination using the area under the receiver operating characteristic curve (AUC) and compared discrimination between the derivation and validation models based on the AUC estimates and overlap of their 95% confidence intervals (CIs). We evaluated whether the association between corticosteroid use and mortality by 30 days from admission differed by latent class by including a corticosteroid-by-class interaction term in logistic regression models restricted to the Uganda cohort, for whom corticosteroid treatment data were available (10).

## RESULTS

Of the 685 patients in the derivation dataset, 467 met inclusion criteria. There were 300 (64.1%) from Uganda, 135 (28.9%) from Rwanda, and 32 (6.8%) from Liberia. Of the 467 patients, 257 (55.0%) were female, the median (IQR) age was 52.0 (37.0-65.0) years, and 128 (27.4%) died within 30 days of admission **(Table 1)**. The most common infections were pulmonary (n=180; 38.5%) and abdominal (n=143; 30.6%), which often occurred in combination with multiple organ dysfunction **(Supplementary Figure 1)**. The most common type of organ dysfunction was cardiovascular (n=349; 74.7%).

**Table 1.** Characteristics and mortality outcomes of the three cohorts of patients with sepsis in Africa included in the pooled derivation dataset: Uganda cohort; Mbarara Regional Referral Hospital (MRRH), 2019-2023; Rwanda cohort, University Teaching Hospital of Kigali (CHUK) and University Teaching Hospital of Butare (CHUB), 2017.

|  | **Total (n=467)** | **Uganda (n=300)** | **Rwanda (n=135)** | **Liberia (n=32)** | **p-value** |
| --- | --- | --- | --- | --- | --- |
| **Demographics and vital signs** |  |  |  |  |  |
| Age (years) | 52.0<br>(37.0–65.0) | 55.0<br>(43.0–66.0) | 42.0<br>(29.0–59.0) | 46.5<br>(30.8–64.2) | <0.001 |
| Female | 257 (55.0%) | 141 (47.0%) | 97 (71.9%) | 19 (59.4%) | <0.001 |
| Temperature (°C) | 37.3<br>(36.4–37.9) | 37.5<br>(36.5–38.0) | 37.0<br>(36.4–38.0) | 36.5<br>(36.4–36.8) | <0.001 |
| Heart rate (bpm) | 124.0<br>(112.0–132.0) | 127.0<br>(121.0–133.0) | 109.0<br>(93.0–124.0) | 110.0<br>(91.5–122.2) | <0.001 |
| Respiratory rate (brpm) | 30.0<br>(24.0–34.0) | 33.0<br>(28.0–37.0) | 24.0<br>(20.0–25.0) | 23.5<br>(22.0–25.8) | <0.001 |
| Systolic blood pressure (mmHg) | 92.0<br>(87.0–98.0) | 90.0<br>(86.0–94.0) | 97.0<br>(90.0–112.5) | 107.5<br>(87.2–147.2) | <0.001 |
| Diastolic blood pressure (mmHg) | 59.0<br>(52.0–64.0) | 57.5<br>(52.0–62.0) | 60.0<br>(52.0–70.0) | 72.0<br>(59.0–87.0) | <0.001 |
| Mean arterial pressure (mmHg) | 69.7<br>(64.0–75.0) | 68.3<br>(63.3–72.3) | 71.3<br>(65.3–83.5) | 82.0<br>(71.9–106.1) | <0.001 |
| Oxygen saturation (%) | 95.0<br>(91.0–97.0) | 94.0<br>(90.0–97.0) | 95.0<br>(93.0–98.0) | 96.5<br>(94.0–97.2) | 0.002 |
| UVA score (n) | 6.0<br>(4.0–8.0) | 9.0<br>(8.0–10.0) | 4.0<br>(3.0–6.0) | 7.0<br>(5.0–8.0) | <0.001 |
| High-risk UVA score | 171 (36.6%) | 87 (29.0%) | 57 (42.2%) | 27 (84.4%) | <0.001 |
| **Infection types** |  |  |  |  |  |
| HIV | 127 (27.2%) | 88 (29.3%) | 33 (24.4%) | 6 (18.8%) | 0.35 |
| Tuberculosis | 39 (8.4%) | 34 (11.3%) | 2 (1.5%) | 3 (9.4%) | <0.001 |
| Pulmonary | 180 (38.5%) | 113 (37.7%) | 60 (44.4%) | 7 (21.9%) | 0.06 |
| Abdominal | 143 (30.6%) | 89 (29.7%) | 49 (36.3%) | 5 (15.6%) | 0.06 |
| Central Nervous System | 66 (14.1%) | 37 (12.3%) | 23 (17.0%) | 6 (18.8%) | 0.27 |
| Urinary | 57 (12.2%) | 42 (14.0%) | 14 (10.4%) | 1 (3.1%) | 0.15 |
| Soft Tissue | 36 (7.7%) | 25 (8.3%) | 11 (8.1%) | 0 (0.0%) | 0.28 |
| Bacteremia | 8 (1.7%) | 5 (1.7%) | 3 (2.2%) | 0 (0.0%) | 0.84 |
| Malaria | 28 (6.0%) | 17 (5.7%) | 0 (0.0%) | 11 (34.4%) | <0.001 |
| Multiple infections | 110 (23.6%) | 68 (22.7%) | 34 (25.2%) | 8 (25.0%) | 0.82 |
| Undifferentiated | 33 (7.1%) | 9 (3.0%) | 10 (7.4%) | 14 (43.8%) | <0.001 |
| **Organ dysfunction** |  |  |  |  |  |
| Cardiovascular | 349 (74.7%) | 263 (87.7%) | 68 (50.4%) | 18 (56.2%) | <0.001 |
| Respiratory | 255 (54.6%) | 221 (73.7%) | 28 (20.7%) | 6 (18.8%) | <0.001 |
| Altered Mental Status | 306 (65.5%) | 231 (77.0%) | 48 (35.6%) | 27 (84.4%) | <0.001 |
| Multiple | 302 (64.7%) | 250 (83.3%) | 36 (26.7%) | 16 (50.0%) | <0.001 |
| **Mortality** |  |  |  |  |  |
| Death by 30 days from admission (%) | 128 (27.4%) | 49 (16.3%) | 61 (45.2%) | 18 (56.2%) | <0.001 |

Data are presented as n (%) or median [interquartile range]; bpm, beats per minute; brpm, breaths per minute; HIV, human immunodeficiency virus; UVA, Universal Vital Assessment.

### Cramér’s V analysis

Associations among infection source and organ dysfunction were similar in survivors and non-survivors **(Supplementary Figure 2)**. Tuberculosis was strongly associated with HIV (Cramér’s V=0.57) and moderately associated with having multiple sites of infection (Cramér’s V=0.40). Neurological dysfunction was strongly associated with CNS infection (Cramér’s V=0.47) and having multiple sites of infection (Cramér’s V=0.51).

### Logistic regression analysis

After controlling for age, sex, and enrollment site (Uganda), participants with a high-risk UVA score (>4), altered mental status, multiple organ dysfunction, CNS infection, living with HIV, and cardiovascular dysfunction were associated with higher odds of mortality than participants without each respective characteristic **(Figure 1)**. Participants with soft tissue infection had lower odds of mortality than those without soft tissue infection. Infection source-organ dysfunction pairs that were associated with higher odds of mortality included CNS infection combined with living with HIV, neurological dysfunction, cardiovascular dysfunction, respiratory dysfunction or multiple organ dysfunction; having multiple sites of infection combined with living with HIV or neurological dysfunction; and pulmonary infection combined with neurological dysfunction **(Supplementary Figure 3)**. Bacteremia combined with any organ dysfunction was associated with higher odds of mortality.

**Figure 1.**
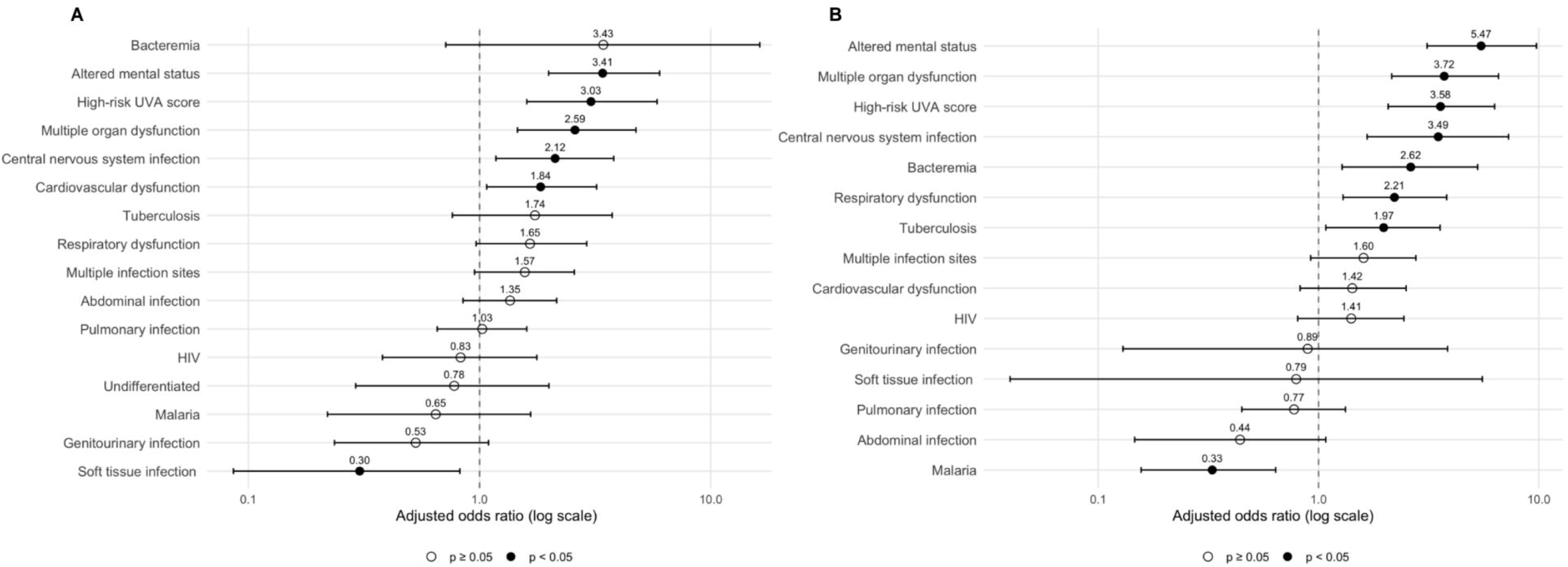
Forest plot of adjusted odds ratios for the associations of infection sites and organ dysfunctions with in-hospital mortality in **(A)** the derivation dataset and **(B)** the validation dataset. Models were adjusted for age, sex, and Uganda enrollment site. Abbreviations: HIV, human immunodeficiency virus; UVA, Universal Vital Assessment.

### Latent class analysis

In the latent class analysis, we selected the four-class model, which had the second-lowest BIC (4521.72) and AIC (4326.84), because it provided the optimal balance of statistical fit, parsimony, and clinically interpretable class structure **(Figure 2)**. This model had a likelihood ratio statistic (G²) of 413.68 and a Pearson chi-square goodness-of-fit statistic of 847.64.

**Figure 2.**
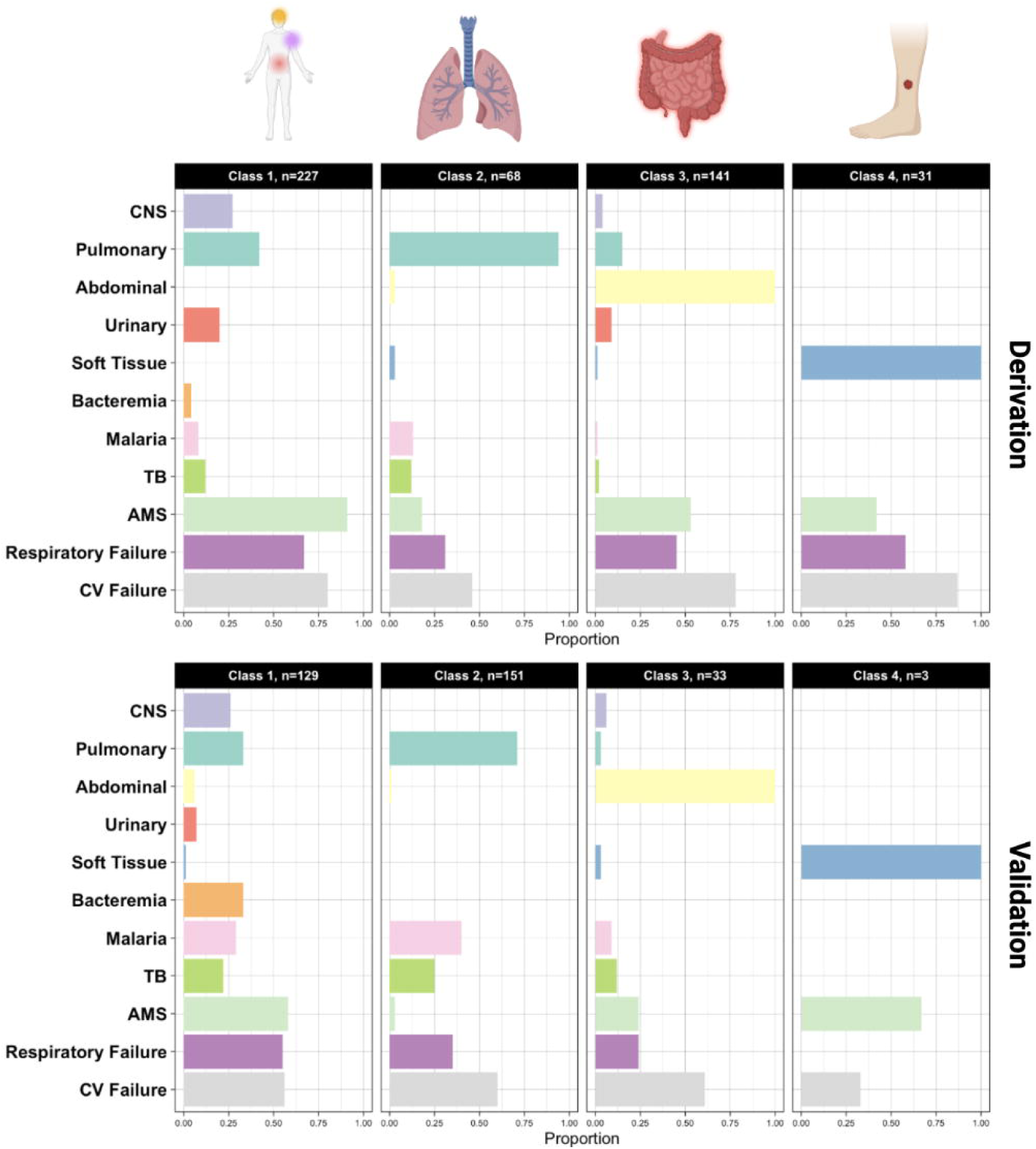
Comparison of latent class profiles between the derivation dataset and the validation dataset after application of the derivation latent class model. Abbreviations: AMS, altered mental status; CNS, central nervous system; CV, cardiovascular; TB, tuberculosis.

Classification performance was excellent, with an entropy of 0.88, indicating clear separation between classes and high confidence in class assignment. Class 1 (n=227) was characterized by heterogeneous infection sources without a single predominant source and neurological, cardiovascular, and respiratory dysfunction, with 30-day mortality of 30.8%; Class 2 (n=68) was characterized by pulmonary infection, with 16.2% mortality; Class 3 (n=141) was characterized by abdominal infection, with 31.9% mortality; and Class 4 (n=31) was characterized by soft tissue infection and cardiovascular dysfunction, with the lowest mortality (6.5%) **(Figure 3)**. Mortality by 30 days from admission differed significantly across the four classes (p=0.003) (Supplementary Figure 4).

**Figure 3.**
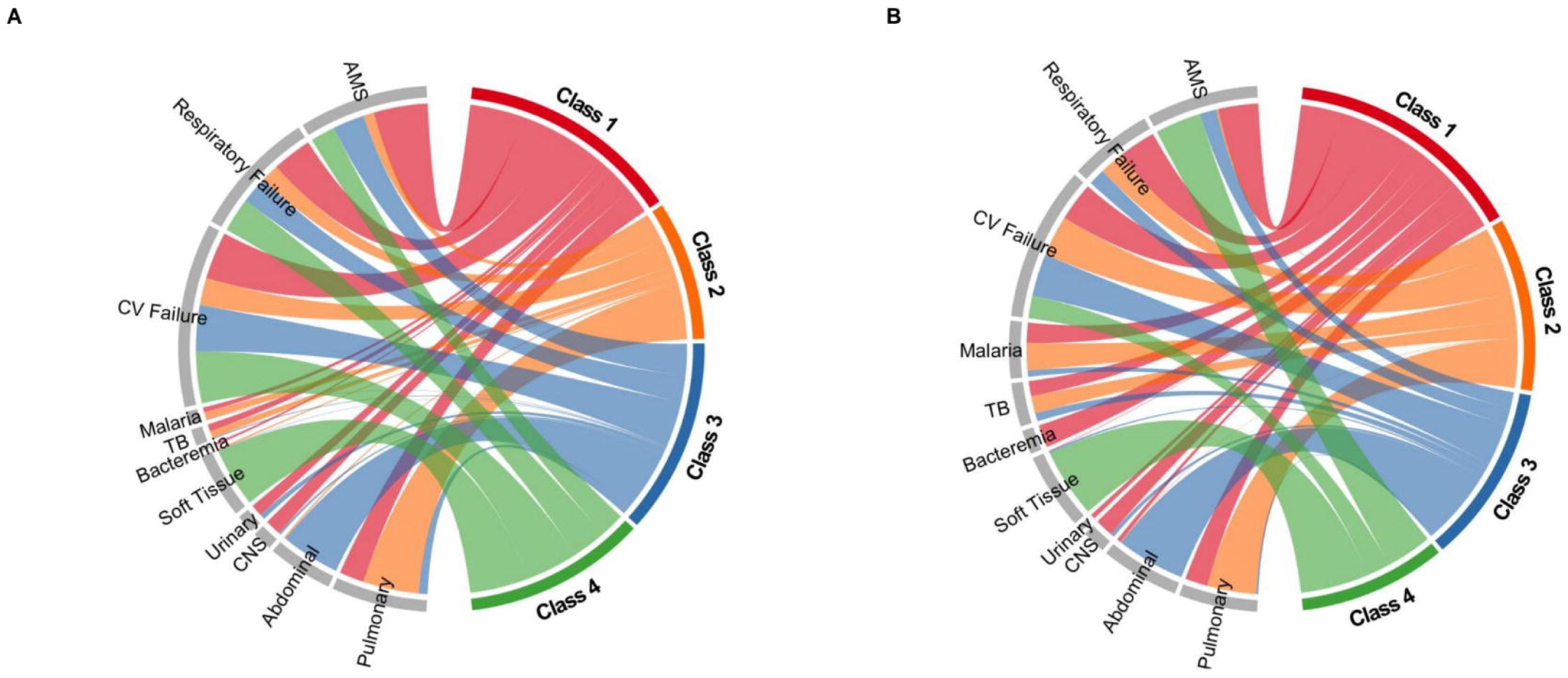
Chord plots illustrating the relative proportions of infection sites and organ dysfunctions to each latent class in **(A)** the derivation dataset and **(B)** the validation dataset following class assignment using the derivation model. Abbreviations: AMS, altered mental status; CNS, central nervous system; CV, cardiovascular; TB, tuberculosis.

In logistic regression analyses comparing membership in each class with membership in all other classes, Classes 1 and 3 were not associated with mortality **(Supplementary Figure 5; Table 2)**; however, membership in Class 2 (odds ratio [OR] 0.45, 95% confidence interval [CI] 0.23-0.90, p=0.03) or Class 4 (OR 0.17, 95% CI 0.04-0.74, p=0.02) was associated with lower odds of mortality. In the multivariable analysis comparing classes simultaneously and using Class 1 as the reference, Classes 2 (OR 0.22, 95% CI 0.10-0.47, p=0.001) and 4 (OR 0.14, 95% CI 0.02-0.52, p=0.01) were associated with lower odds of mortality, while mortality in Class 3 did not differ significantly from Class 1 (OR 0.90, 95% CI 0.54-1.48, p=0.67) **(Figure 4)**. The model demonstrated good discrimination for 30-day mortality (AUC=0.72, 95% CI 0.67-0.78).

**Figure 4.**
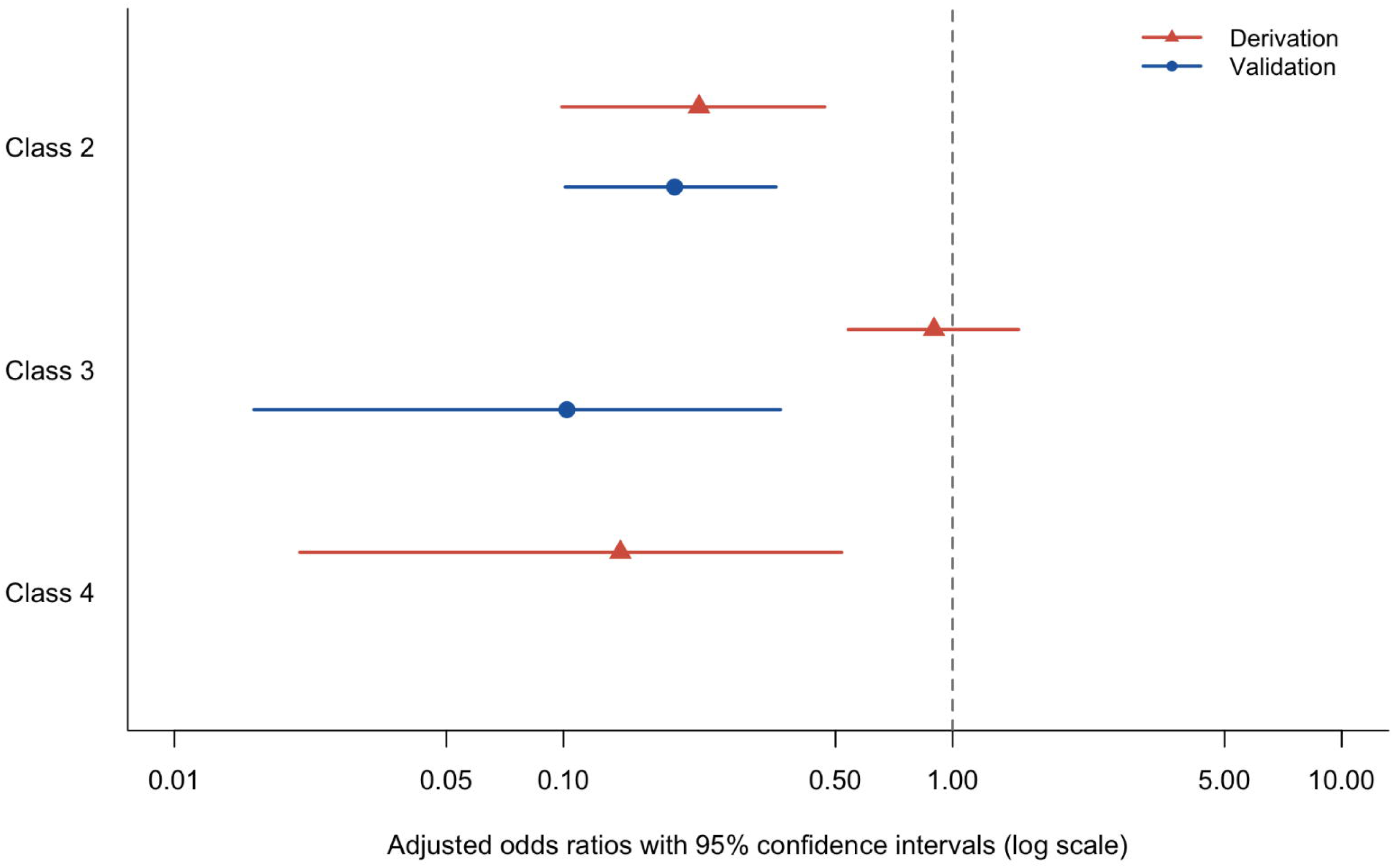
Forest plot of adjusted odds ratios for in-hospital mortality comparing latent classes with Class 1 (heterogeneous-source infection subphenotype) as the reference in the derivation and validation datasets. Models were adjusted for age, sex, and enrollment site (Uganda, derivation cohort only). Class 4 was excluded from the validation dataset analysis because of its small sample size.

**Table 2.** Estimated odds ratios for 30-day mortality obtained from multivariable logistic regression adjusted for age, sex, and enrollment site (Uganda) in the derivation cohort. The first analysis compared each latent class with all other participants. The second analysis compared latent classes simultaneously, with Class 1 (heterogeneous-source infection subphenotype) serving as the reference group.

|  | **Adjusted odds ratio** | **95% Confidence interval** | **p-value** |
| --- | --- | --- | --- |
| Class 1 | 3.87 | (1.79-8.37) | 0.0006 |
| Class 2 | 0.99 | (0.58-1.71) | 0.98 |
| Class 3 | 1.12 | (0.61-2.07) | 0.72 |
| Class 4 | 0.33 | (0.15–0.73) | 0.006 |
|  | **Adjusted odds ratio** | **95% Confidence interval** | **p-value** |
| Class 1 |  | Referent |  |
| Class 2 | 0.30 | (0.13-0.70) | 0.005 |
| Class 3 | 0.33 | (0.14-0.81) | 0.02 |
| Class 4 | 0.12 | (0.04-0.34) | 0.0001 |

We found no evidence that the association between corticosteroid therapy and mortality by 30 days from admission differed across latent classes in the Uganda cohort **(Supplementary Table 3)**.

### Validation dataset

Of the 1,473 patients in the validation dataset, 316 met inclusion criteria among whom 184 (58.5%) were female, the median (IQR) age was 35.7 (17.0-95.0) years, and 128 (40.5%) died **(Supplementary Table 4)**. The most common infections were pulmonary (n=150; 47.5%) and malaria (n=101; 32.0%). The most common type of organ dysfunction was cardiovascular (n=183; 57.9%). Associations of infection source and organ dysfunction with mortality by 30 days from admission were generally directionally consistent and with a similar magnitude compared with those observed in the derivation dataset **(Figure 1)**.

When applied to the validation dataset, the derived latent class model demonstrated high classification certainty, with mean maximum posterior probabilities ranging from 0.89 to 0.92 and an entropy of 0.78 **(Supplementary Figure 6; Supplementary Table 5)**. The model reproduced four corresponding subphenotypes **(Figures 2 and 3)**. Class 1 had the highest mortality (39.5%), followed by Class 2 (11.3%) and Class 3 (6.1%). Class 4 included only three patients and was too small for reliable comparison **(Supplementary Figure 4)**.

In logistic regression analyses comparing membership in each class with membership in all other classes, Class 1 was associated with higher odds of mortality (OR 5.46, 95% CI 3.03-9.84, p<0.001), while Classes 2 (OR 0.26, 95% CI 0.14-0.47, p<0.001) and 3 (OR 0.21, 95% CI 0.05-0.90, p=0.04) were associated with lower odds of mortality **(Supplementary Figure 5; Supplementary Table 6)**. In the multivariable analysis comparing classes simultaneously and using Class 1 as the reference, Classes 2 (OR 0.19, 95% CI 0.10-0.35, p<0.001) and 3 (OR 0.10, 95% CI 0.02-0.36, p=0.003) were associated with lower odds of mortaliy **(Figure 4; Supplementary Table 6)**. The model demonstrated good discrimination (AUC 0.73, 95% CI 0.67-0.80), with overlapping 95% CIs for the AUC estimates in the derivation and validation datasets.

## DISCUSSION

In this analysis of pooled cohorts of adults with sepsis from Africa, we observed substantial heterogeneity in the combinations of infection sites and organ dysfunction. Using the pooled derivation dataset, we identified four distinct clinical subphenotypes that were primarily defined by infection site and associated with different mortality risks. Application of the latent class model derived from this dataset to an independent validation dataset yielded reproducible classification and similar mortality patterns. These findings suggest that the identified subphenotypes are reproducible across diverse African patient populations. These clinically defined subphenotypes could provide a pragmatic approach to risk stratification, triage, and patient monitoring in resource-limited settings in Africa.

Our previous study of adults hospitalized with sepsis in Uganda demonstrated that mortality increased with the number of organ dysfunctions present, irrespective of the presence of septic shock (18). However, that analysis did not account for infection source or the specific pattern of organ dysfunction. Consistent with our current findings, a large multicenter study from the United States also found that mortality varied according to both infection source and organ dysfunction, with the highest mortality observed among patients with bacteremia and respiratory failure requiring mechanical ventilation (19). Importantly, mortality was not determined solely by the number of organ dysfunctions but also by the specific combination of infection source and organ dysfunction. By integrating these clinical features using latent class analysis, our study extends these observations by identifying reproducible clinical subphenotypes with distinct mortality risks that were validated across independent African cohorts.

Our findings are broadly consistent with prior studies that have identified reproducible sepsis clinical subphenotypes using unsupervised learning. In the Sepsis Endotyping in Emergency Care (SENECA) study that used clinical data from the United States, latent class analysis identified four distinct sepsis clinical subphenotypes (α, β, γ, and δ) characterized by differences in inflammation, organ dysfunction, comorbidities, and mortality (7). The α subphenotype exhibited the lowest illness severity and mortality, whereas the δ subphenotype was characterized by septic shock, liver dysfunction, and the highest mortality. However, the SENECA subphenotypes were derived using a broader set of clinical and laboratory variables than were routinely available in our cohorts and therefore could not be directly applied to our data. Instead, we derived and externally validated four clinical subphenotypes based on infection source and organ dysfunction. We retained Classes 1-4 as the primary class designations and used descriptive labels based on their predominant clinical features to facilitate interpretation. Although the defining characteristics differ from those reported in SENECA, both studies demonstrate that clinically derived subphenotypes capture meaningful heterogeneity in sepsis and identify patient groups with markedly different prognoses.

The observed clustering of infection source and organ dysfunction is biologically plausible in the context of organ crosstalk during sepsis (20). Organ dysfunction results from bidirectional communication among organs through inflammatory, neurohormonal, metabolic, and immune pathways rather than from isolated organ injury (21). Sepsis-induced injury to one organ can propagate secondary dysfunction in distant organs, creating characteristic patterns of multiorgan injury (22, 23). For example, pulmonary infections may promote respiratory, cardiovascular, and renal dysfunction, whereas central nervous system infections are often accompanied by neurological dysfunction (24–26). These interconnected mechanisms provide a biological framework for the reproducible subphenotypes identified in our study and suggest that clinically observed patterns of infection and organ dysfunction reflect underlying pathophysiologic networks.

In our study, patients in the Class 1 (heterogeneous-source infection) subphenotype had the highest risk of mortality, whereas those in the Class 4 (soft tissue infection) subphenotype had the lowest. The high mortality observed in the Class 1 subphenotype likely reflects a combination of severe organ dysfunction and the inability to localize an infectious source at presentation, which may indicate greater diagnostic uncertainty or a more advanced systemic disease process. Consistent with this finding, a large multicenter study from the United States reported that, after adjustment, patients with blood culture-positive sepsis had lower odds of mortality than those with culture-negative sepsis despite similar crude mortality rates, suggesting that failure to identify an infectious source may limit opportunities for targeted therapy (27). In contrast, patients in the Class 4 subphenotype in our study generally had less severe organ dysfunction and a readily identifiable and treatable source of infection. Together, these findings suggest that infection source, in addition to organ dysfunction, is an important determinant of prognosis and supports the use of clinically defined subphenotypes for early risk stratification.

Although these subphenotypes identified patients with markedly different mortality risks, we found no evidence that latent class modified the association between corticosteroid therapy and mortality by 30 days from admission. Our subphenotypes were based on infection source and organ dysfunction rather than host inflammatory or immunologic profiles that may better predict response to immunomodulatory therapies. In addition, we evaluated corticosteroid treatment only in the Uganda cohort, and treatment was administered as part of routine clinical care rather than within a clinical trial. These findings suggest that clinically defined subphenotypes are prognostic but may not alone predict response to corticosteroid therapy. Future studies that integrate clinical subphenotypes with host-response biomarkers or molecular endotypes may better identify treatment-responsive patient groups and should be evaluated prospectively in randomized clinical trials.

This study has limitations. First, the retrospective design is subject to information bias, residual confounding, and potential misclassification. Eligibility required a clinical diagnosis of infection and ≥2 qSOFA criteria. Because qSOFA has recognized limitations as a standalone screening tool for sepsis and overlaps with the organ dysfunction variables used in the latent class analysis, our inclusion criteria may have selected for specific patterns of organ dysfunction and limited representation of the broader sepsis population. Second, the relatively small sample size for some infection sources may have limited statistical power and reduced the ability of latent class analysis to identify additional clinically meaningful subphenotypes. Third, variability in data completeness and documentation across cohorts may have affected classification of infection source and organ dysfunction, although we mitigated the impact of missing data through imputation. In addition, our analyses relied on clinical variables measured at a single time point and did not capture the temporal evolution of organ dysfunction, changes in vital signs, sepsis recognition, or the development of concurrent infections over the course of illness. Fourth, differences in infection source distributions, HIV prevalence, pathogen distributions, and other unmeasured site-specific factors across cohorts may limit generalizability and influence observed outcomes. Several infection phenotypes were less common in the validation dataset, resulting in small Classes 3 and 4 and limiting statistical power and precision. Nevertheless, successful external validation across cohorts with differing epidemiologic characteristics supports the reproducibility of the identified subphenotypes across diverse African settings.

Finally, the derived subphenotypes were based on routinely available clinical variables and were not compared with biologic or molecular endotypes. Future studies integrating clinical, biomarker, and multi-omic data may further refine these clinically defined subphenotypes and provide additional insight into their underlying mechanisms.

In conclusion, in this pooled analysis of adults hospitalized with sepsis in Africa, latent class analysis identified four reproducible clinical subphenotypes defined primarily by infection source. These subphenotypes were associated with distinct mortality risks and were externally validated in an independent dataset, demonstrating that routinely available clinical features can identify clinically meaningful sepsis subgroups. These findings provide a practical framework for risk stratification and may inform future subphenotype-guided clinical trials and targeted treatment strategies. Prospective studies are needed to further validate these subphenotypes and determine whether subphenotype-guided management improves patient outcomes.

## Data Availability

All data produced in the present study are available upon reasonable request to the authors.

---

**Supplementary Figure 1.** UpSet plot showing combinations of infection sites and organ dysfunctions among adults with sepsis in the pooled derivation dataset. Abbreviations: AMS, altered mental status; CNS, central nervous system; CV, cardiovascular; HIV, human immunodeficiency virus; TB, tuberculosis.

**Supplementary Figure 2.** Cramér’s V analysis showing the strength of association between infection sites and organ dysfunctions in **(A)** the entire population, **(B)** survivors, and **(C)** non-survivors in the derivation dataset. Abbreviations: HIV, human immunodeficiency virus.

**Supplementary Figure 3.** Heat map of adjusted odds ratios for infection site-organ dysfunction pairs as predictors of in-hospital mortality in the derivation dataset. Models were adjusted for age, sex, and enrollment site (Uganda).

**Supplementary Figure 4.** Comparison of percent mortality by 30 days from admission across latent classes in the derivation and validation datasets.

**Supplementary Figure 5.** Comparison of adjusted odds ratios for percent mortality by 30 days from admission for each latent class versus all other participants in either the derivation or validation dataset. Separate multivariable logistic regression models were adjusted for age, sex, and enrollment site (Uganda, derivation cohort only).

**Supplementary Figure 6.** Mean posterior probability of class membership by assigned latent class in the validation dataset.

