## Supplementary material for "Derivation and validation of clinical subphenotypes of adult sepsis in Africa: A latent class analysis of infection site and organ dysfunction"

| **Table of Contents** | |
| --- | --- |
| **Supplementary Table** | **Page** |
| Table S1 | 3 |
| Table S2 | 4 |
| Figure S1 | 5 |
| Figure S2 | 6 |
| Figure S3 | 7 |
| Figure S4 | 8 |
| Figure S5 | 9 |
| Figure S6 | 10 |

**Supplementary Table 1.** Categorization of infection site based on collected case definitions from each cohort study.

| Infection site | Uganda | Rwanda | Liberia | Documented diagnoses |
| --- | --- | --- | --- | --- |
| Pulmonary | Pneumonia | Lung, pulmonary | Respiratory system | Pneumonia, lung abscess |
| Abdominal | Peritonitis, enteritis, IAI | Abdominal | Gastrointestinal | Enteritis, peritonitis, IAI |
| CNS | Meningitis | CNS | CNS | Meningitis, encephalitis |
| Genitourinary | Genitourinary | Urine | Genitourinary | UTI, pyelonephritis |
| Soft Tissue | Cellulitis | Skin/wound, Joint | Soft tissue infection | Cellulitis, osteomyelitis, wound infection, septic arthritis |
| Bacteremia |  | Heart/Heart valves |  | Endocarditis, septicemia, bacteremia, toxic shock syndrome |
| Malaria | N/A | Malaria | Malaria | Malaria |
| Tuberculosis | clinically or previously diagnosed | positive culture | clinically or previously diagnosed | Tuberculosis; pulmonary, meningitis, disseminated |
| Undifferentiated | Other/Unknown, with unspecified diagnosis or noninfectious diagnosis | | | |

CNS, central nervous system; IAI, intra-abdominal infection, N/A, not applicable; UTI, urinary tract infection.

**Supplementary Table 2.** Criteria for organ dysfunction criteria and clinical scores.

| Organ dysfunction | Criteria |
| --- | --- |
| Cardiovascular dysfunction (any of the following): | Heart rate ≥ 120 bpm  Systolic blood pressure < 90 mmHg  Mean arterial pressure < 65 mmHg  Received vasopressors |
| Respiratory dysfunction (any of the following) | Respiratory rate ≥ 30 brpm  Oxygen saturation < 90% |
| Altered mental status | Glasgow coma scale score < 15 |
| Quick sequential organ failure assessment score (≥2) | Respiration rate ≥ 22 brpm  Systolic blood pressure ≥100 mmHg  Glasgow coma scale score < 15 |
| Universal Vital Assessment score (<2, low-risk; 2-4, medium-risk; >4, high-risk) | Temperature <35°C: 2 points  Heart Rate ≥ 120 bpm: 1 point  Respiratory Rate ≥ 30 breaths/min: 1 point  Systolic blood pressure <90 mmHg: 1 point  Oxygen Saturation < 90%: 2 points  Glasgow coma scale score <15: 4 points  HIV positive: 1 point |

Bpm, beats per minute; brpm, breaths per minute; HIV, human immunodeficiency virus

**Supplementary Figure 1**

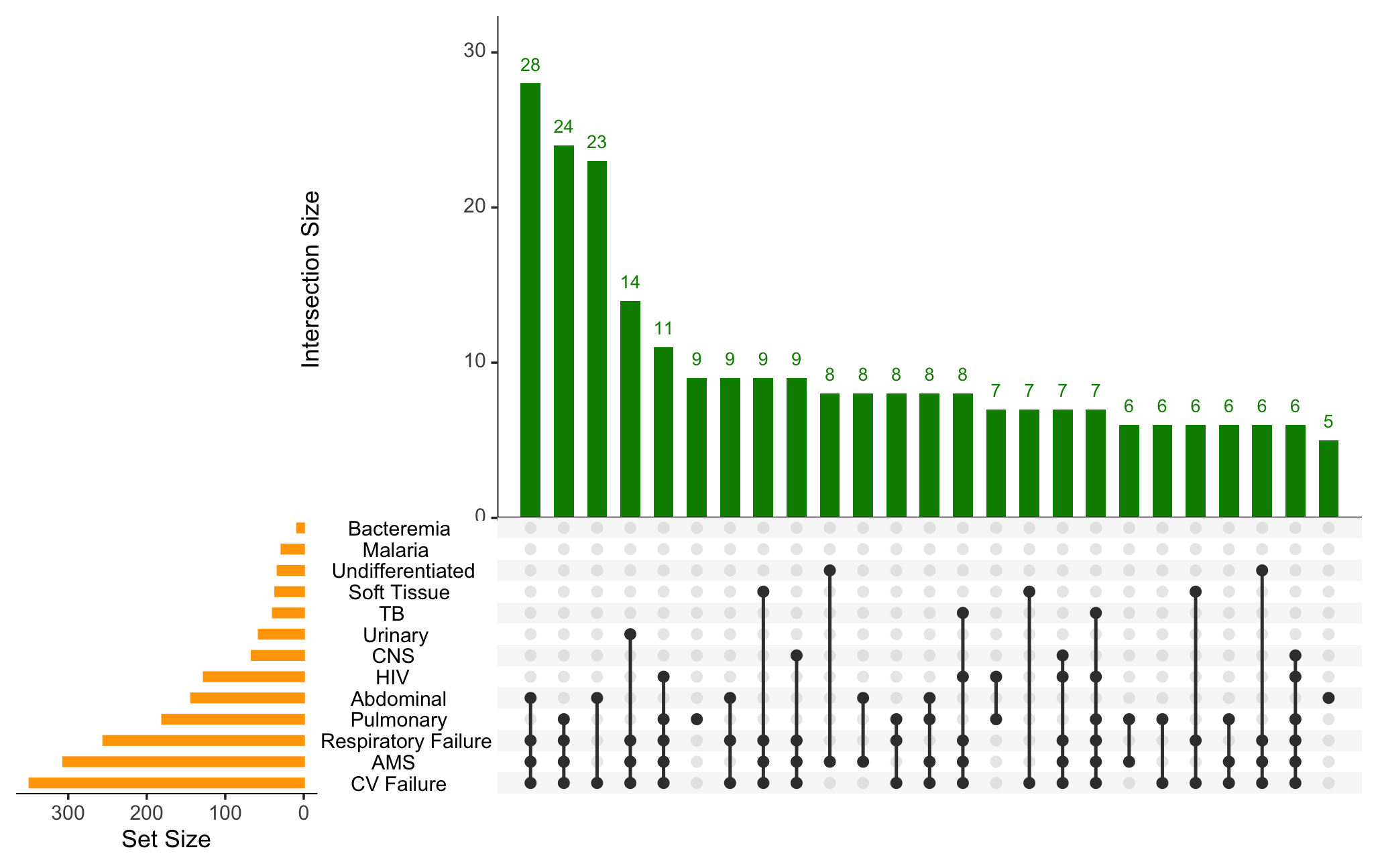

**Supplementary Figure 2**

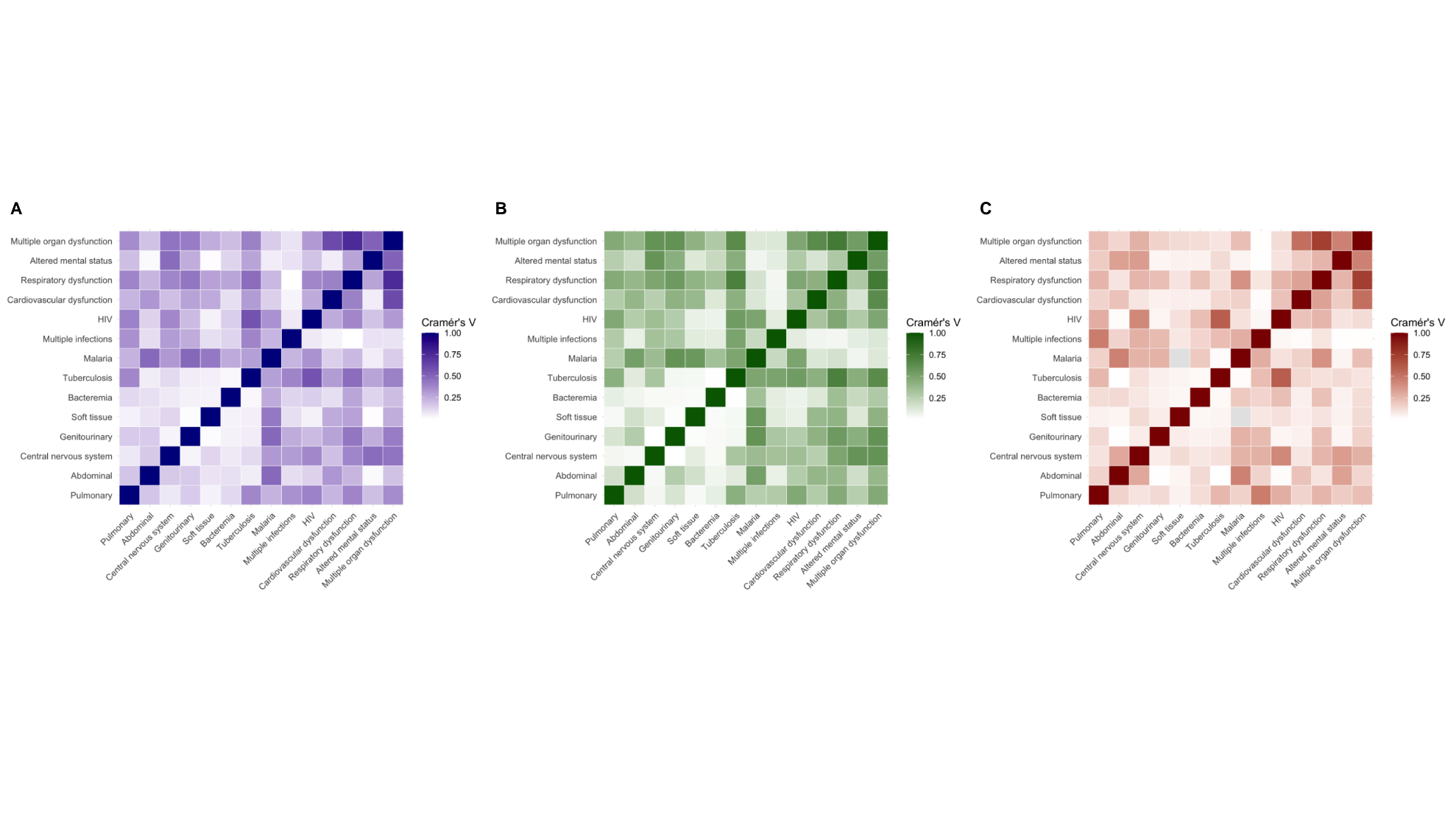

**Supplementary Figure 3**

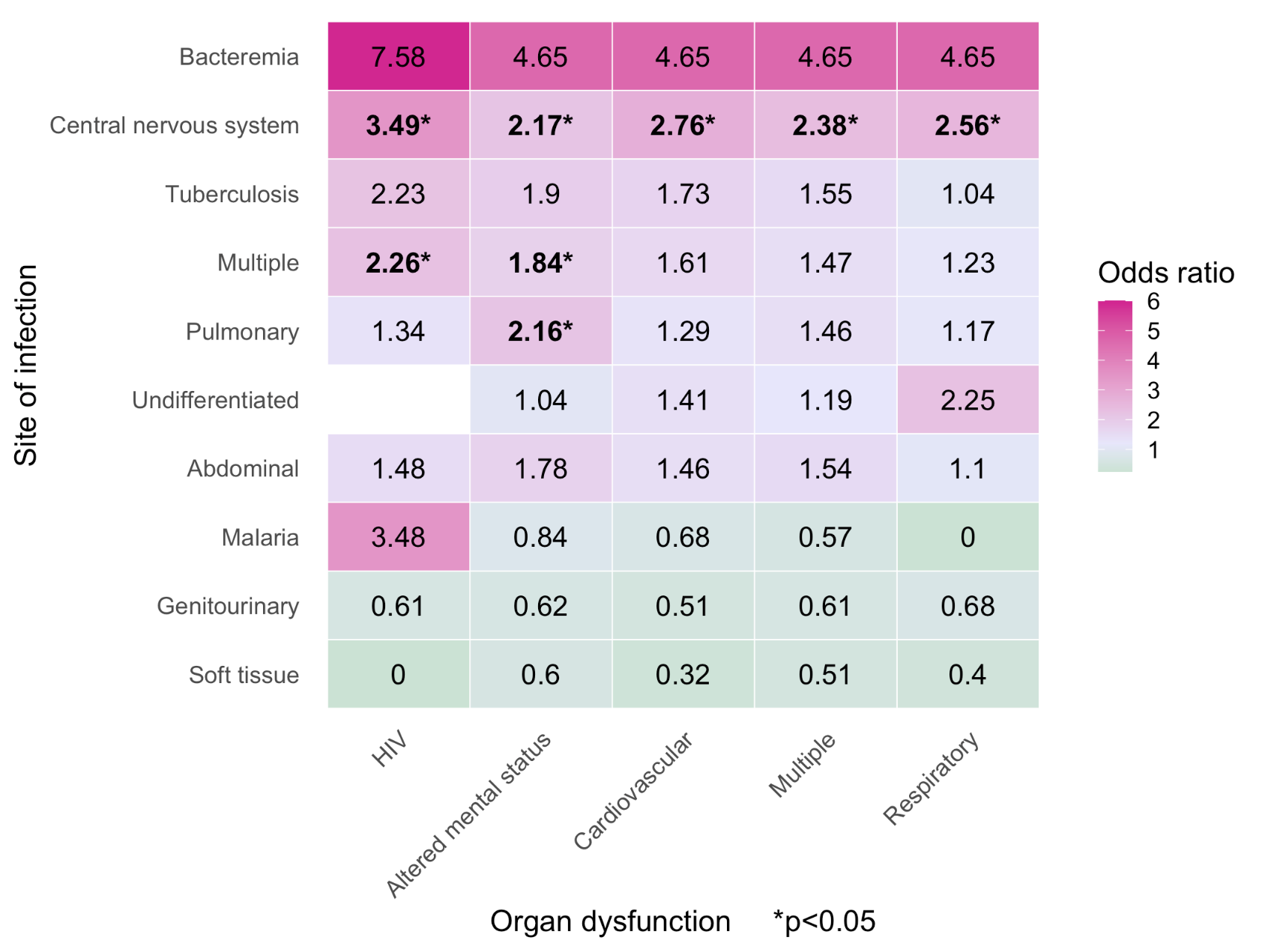

**Supplementary Figure 4**

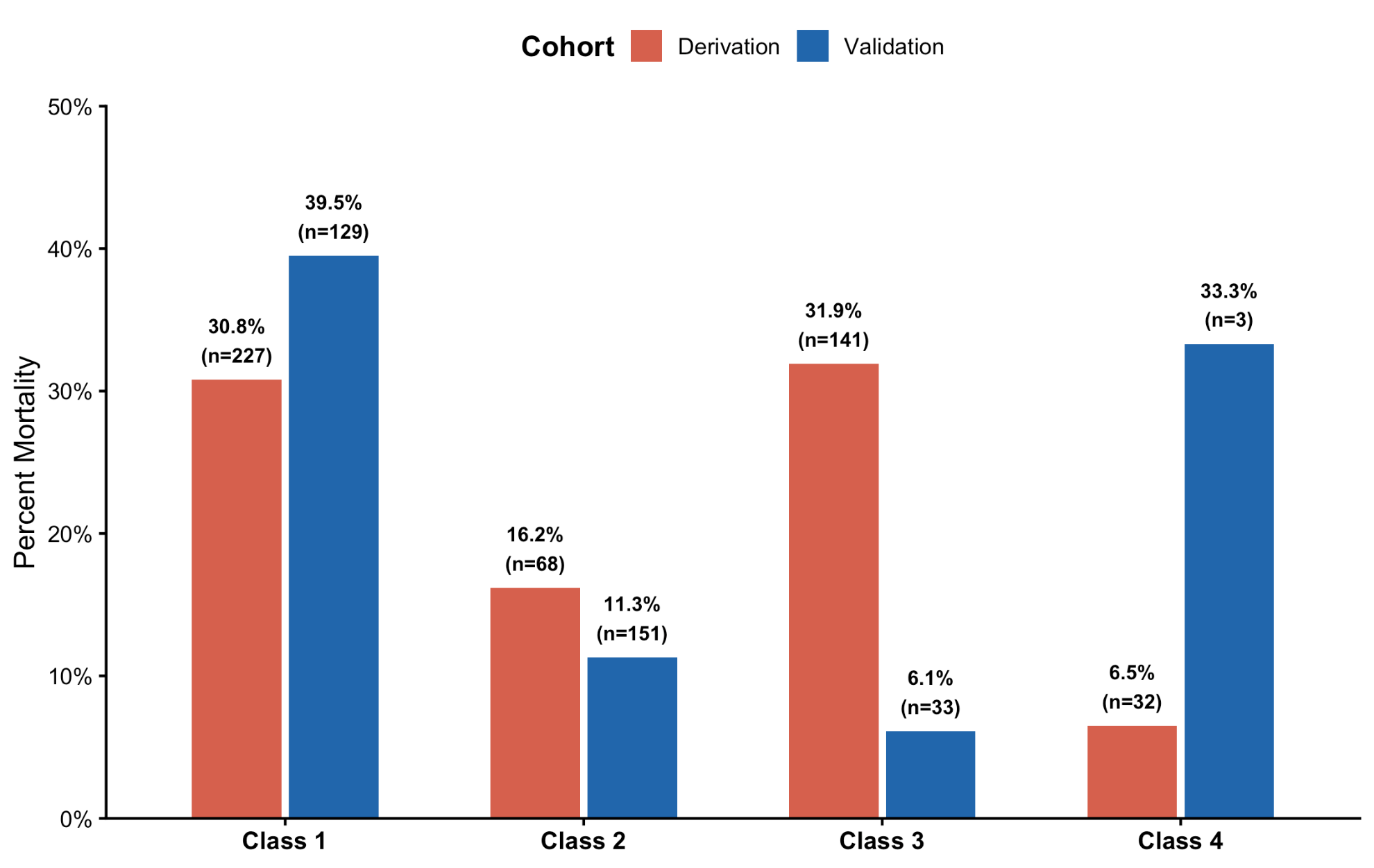

**Supplementary Figure 5**

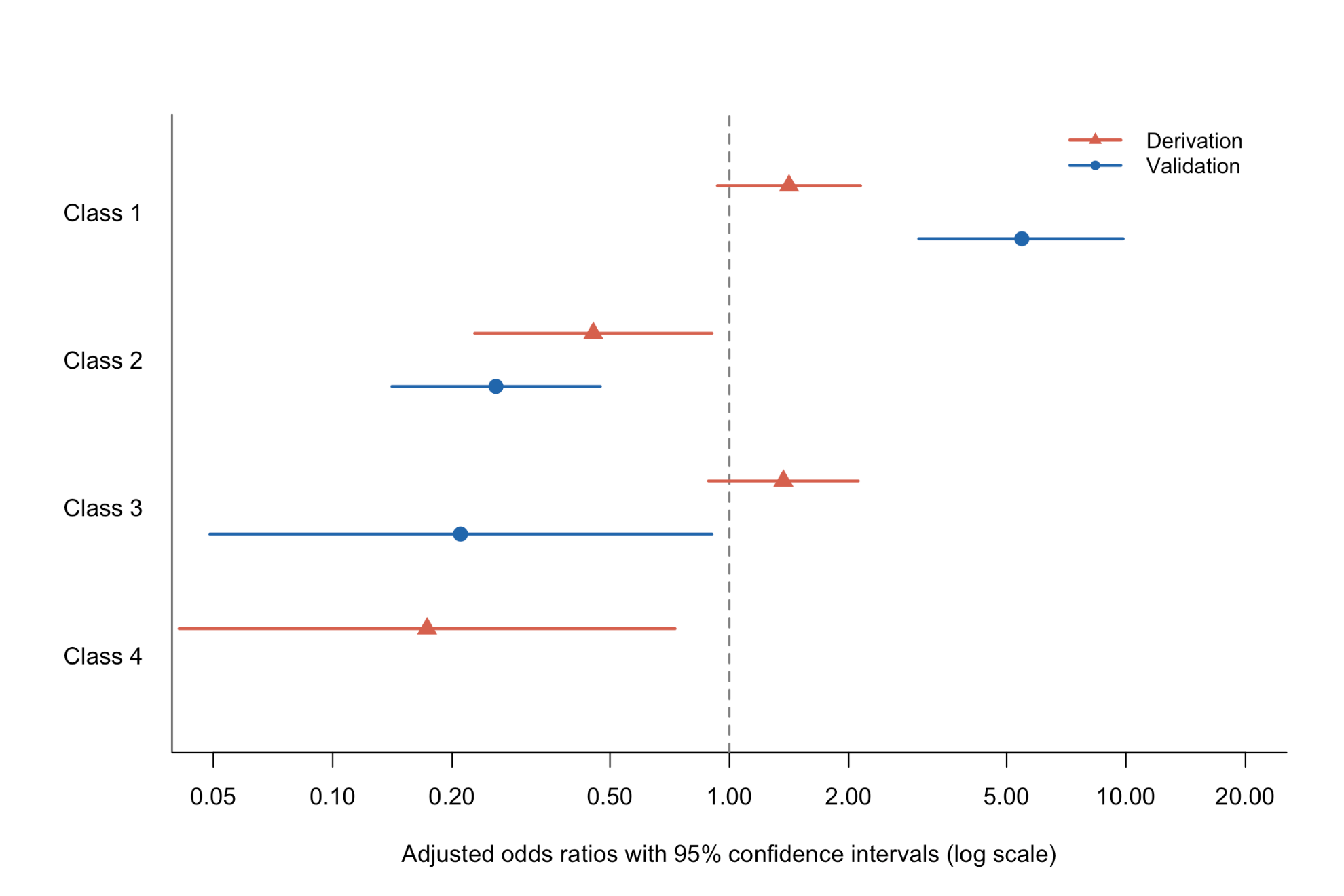

**Supplementary Figure 6**

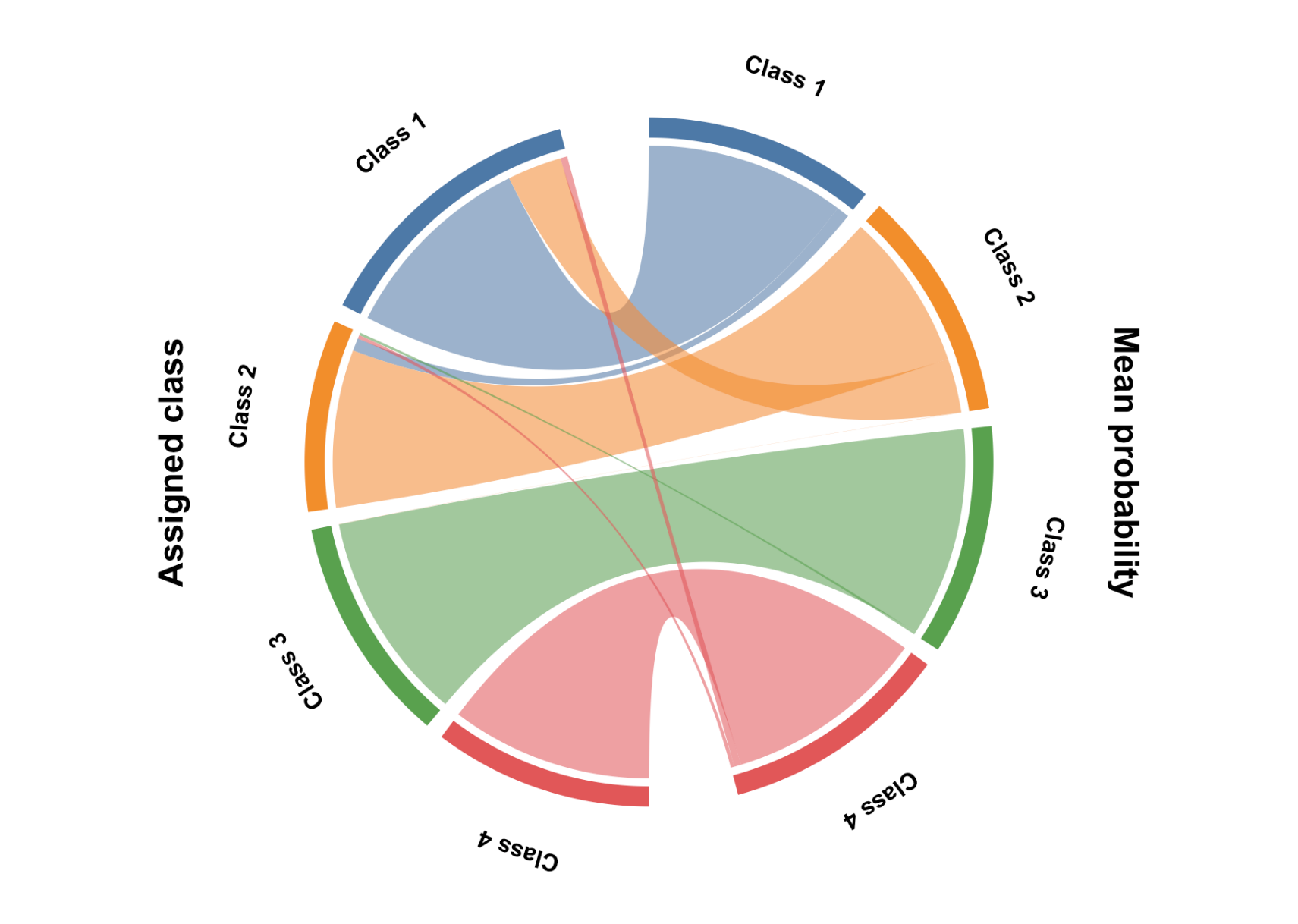
